# Clinical improvement of DM1 patients reflected by reversal of disease-induced gene expression in blood

**DOI:** 10.1101/2022.03.11.22272021

**Authors:** Remco T.P. van Cruchten, Daniël van As, Jeffrey C. Glennon, Baziel G.M. van Engelen, Peter A. C. ’t Hoen, the OPTIMISTIC consortium, the ReCognitION consortium

**Author notes:** **Corresponding author** Professor dr. P.A.C. ‘t Hoen, Centre for Molecular and Biomolecular Informatics, Radboud University Medical Center, Nijmegen, The Netherlands. Shared first authors, contributed equally (Remco T.P. van Cruchten, Daniël van As). Shared senior authors, contributed equally (Baziel G.M. van Engelen, Peter A. C. ’t Hoen). Several authors of this publication are members of the Radboudumc Center of Expertise for neuromuscular disorders (Radboud-NMD), Netherlands Neuromuscular Center (NL-NMD) and the European Reference Network for rare neuromuscular diseases (EURO-NMD).

## Abstract

Myotonic dystrophy type 1 (DM1) is an incurable multisystem disease caused by a CTG-repeat expansion in the DM1 protein kinase (*DMPK*) gene. The OPTIMISTIC clinical trial demonstrated positive and heterogenous effects of cognitive behavioral therapy (CBT) on the capacity for activity and social participations in DM1 patients. Here, we performed mRNA sequencing of full blood for 27 patients of the OPTIMISTIC cohort before and after the CBT intervention. We identified 608 genes for which their expression was significantly associated with the disease causing CTG-repeat expansion, as well as 1176 genes significantly associated with the average clinical response towards the intervention. Remarkably, all 97 genes significantly associated with both returned to more normal levels in patients who benefited most from CBT. This trend was consistent with the difference observed between DM1 patients and controls in an earlier study of blood mRNA expression levels, singling these genes out as candidate biomarkers for therapy response. Together these results highlight the ability to find disease relevant information in full blood of DM1 patients, opening new avenues to monitor therapy effects.

## 1. Introduction

Myotonic dystrophy type 1 (DM1) is a neuromuscular disease with a worldwide average prevalence of around 1 in 8,000 people and a high unmet clinical need [1]. DM1 is considered the most frequently occurring adult-onset form of muscular dystrophy. This degenerative multisystem disease is characterized by a wide range of symptoms including myotonia, muscle weakness and dystrophy, fatigue, apathy, cataracts, obesity and insulin resistance. Next to a severe decrease of life quality, DM1 patients suffer from a reduced life expectancy mostly due to problems with cardiac and respiratory function. Currently no curative therapy exists.

DM1 is caused by expansion of a CTG trinucleotide microsatellite repeat in the 3’ UTR of the DM1 protein kinase (*DMPK*) gene [2–4]. Unaffected individuals carry up to approximately 37 CTG triplets in *DMPK*, whilst in DM1 patients this ranges from 50 to even a few thousand repetitions. Depending on the inherited repeat length, DM1 can become manifest at birth or early in life but more frequently becomes apparent in adulthood [1]. In general, the disease manifestation is earlier and more severe with longer repeat expansions. Interruption of the CTG repeat by variants such as CCG or CGG are associated with milder symptoms [5]. The expanded CTG repeat is thought to cause disease mainly via an mRNA gain-of-function mechanism, in which aberrant hairpin structures formed by long CUG repeats are central [6– 9]. Directly or indirectly, these hairpin structures dysregulate the function of RNA binding proteins from the muscleblind-like (MBNL) and CUGBP Elav-like (CELF) families, leading to widespread disturbed RNA processing and consequently altered functions of various proteins [10–12]. Although proven to be the disease causing mutation, clinical symptoms of DM1 are only moderately associated with the CTG repeat or the dysregulation of specific proteins which suggests an involvement of other mechanisms in symptom expression [13–16].

While there are many promising therapeutic oligonucleotides, small molecule drugs and gene therapies in the (pre)clinical pipeline for some of the signs and symptoms of DM1, none are expected to reach widespread clinical application soon. Physical training and increasing activity are currently being applied to relieve DM1 symptoms with marked improvements in relatively mildly affected DM1 patients [17,18], which has furthermore shown to induce biochemical responses in DM1 mouse models [19,20].

The to date largest clinical trial in DM1 was OPTIMISTIC: Observational Prolonged Trial In Myotonic dystrophy type 1 to Improve Quality of Life-Standards, a Target Identification Collaboration [18]. The OPTIMISTIC clinical trial included over 250 well-characterized DM1 patients from four centers in Europe, where the effects of cognitive behavioural therapy (CBT) and optional graded exercise therapy were closely monitored over 16 months via more than twenty outcome measures. Notably, the CBT intervention was tailored towards the specific needs of the patient in a shared decision making process between patient and psychotherapist, allowing for a personalized intervention. The trial has shown significant, yet heterogenous improvements for various signs and symptoms, as well as the capacity for social activity and participation in DM1 [21].

Here, we set out to find molecular profiles associated with the disease causing CTG repeat and therapy response based on full blood mRNA sequencing before and after the CBT intervention of 27 carefully selected patients from the OPTIMISTIC cohort. Given the accessibility of peripheral blood, it has increasingly been used for the successful identification of disease biomarkers for a variety of neurological and psychiatric disorders such as Duchenne Muscular Dystrophy, Huntington’s disease, Major Depressive Disorder and DM1 [22–25].

Furthermore, the multisystem nature of DM1 is known to be reflected by various laboratory abnormalities of blood samples, supporting the relevance of peripheral blood for the identification of disease relevant information [26]. We analyzed gene expression levels as a function of CTG repeat size (as a proxy for disease load/severity) and of the therapy response. Next, we combined these findings and compared the results to various previously published datasets. We were able to identify 608 genes significantly associated with the CTG repeat and further illustrate that 97 of these genes returned towards more normal expression levels in clinical CBT responders.

## 2.0 Methods

### 2.1 Ethical committee approval

The OPTIMISTIC study (NCT02118779) was conducted in accordance with the declaration of Helsinki and approved by the medical-ethical scientific committee for human research at each of the four participating clinical centers. Prior to participation in the OPTIMISTIC study, all participants had provided written informed consent, including a consent for the use of their pseudo anonymized blood, urine and DNA samples [18]. Researchers of this study were not able to link the pseudo anonymized samples to patient identities.

### 2.2 Sample source and patient selection

Samples and metadata used for this study were all gathered during the OPTIMISTIC clinical trial [18]. At the different time points in the trial blood was drawn and a wide range of clinical outcome measures were recorded. Figure one has been generated to illustrate the heterogeneity in changes across all grouped outcome measures, with annotation of all individual outcome measures in the legend.

Patients were selected to represent a balanced subset of the OPTIMITSTIC intervention group (n=128) in order to maximize the generalizability of the study findings. Additionally, patients were selected to represent a continuous uniform distribution of therapy response, as assessed by the primary clinical trial outcome the DM1-Activ-c questionnaire [27]. Finally, we selected for patients that were most completely characterized to facilitate future research. To achieve this, patients were selected for which the DM1-Activ-c questionnaire results were available at each time point (n=104), with less than 20% missing values for other outcome measures (n=81) and without a variant CTG repeat (n=74). Homogeneity of baseline disease severity was accounted for by selecting patients that were within one interquartile range (IQR) of the mean for the baseline variables DM1-Activ-c, 6MWT and CTG repeat length (n=45). One patient was excluded because of polypharmacy (n=44). One patient was excluded because of a drop of 57 points of the DM1-Activ-c score between the baseline and 5 month assessments with a subsequent increase of 55 points between the 5 and 10 month assessments (n=43).

For the final selection we aimed for a continuous uniform distribution of both intervention sites and changes in DM1-Activ-c scores after 10 months. The distribution of the 43 patients over the clinical sites A, B, C and D was: A 12, B 11, C 16, D 4. Therefore, it was decided to select all patients from site D. For the remaining 39 patients, patients were randomly selected as long as each selected patient had a unique change in DM1-Activ-c score. Since only 24 unique delta-DM1-Activ-c scores were left, a second round of random selection was performed allowing each delta-DM1-Activ-c score to be present twice. This process of random selection was repeated until a reasonable site and delta-DM1-Activ-c distribution was achieved, defined as more than 7 patients for site A, B and C, resulting in the final selection of 30 patients. Due to unavailability of samples and unsuccessful RNA sequencing, three patients were later excluded (n=27). The final selection featured a site distribution of 5 times center A, 8 B, 10 C, 4 D, and 22 unique changes DM1-Activ-c scores, with no change in DM1-Activ-c score being present more than twice.

### 2.3 RNA-sequencing

Blood drawn during the OPTIMISTIC trial was collected in Tempus tubes and centrally stored at the New Castle MRC Centre for Rare & Neuromuscular Diseases biobank with strict SOPs and temperature control (−80°C). RNA was locally isolated in Nijmegen using the Tempus Spin RNA Isolation Kit (Applied Biosystems/Thermo Fisher Scientific) according to manufacturer’s instructions. The concentration and RNA Integrity Number (RIN) was checked using Fragment Analyzer (Thermo Fisher Scientific). The mean RIN value was 8.9 and all were > 7.5. Hemoglobin mRNA was depleted using the Globinclear kit (Thermo Fisher Scientific). Libraries were prepared using NEBNext Ultra II Directional RNA Library Prep Kit (Illumina) according to manufacturer’s instructions for a polyA mRNA workflow using UMI-indexed adapters. The size distribution (between 300 and 500 bp) was confirmed using Fragment Analyzer. 150 bp paired end sequencing was performed with a NovaSeq6000 machine (Illumina) at a library concentration of 1.1 nM, generating >30 M read pairs per sample. All raw sequencing data and associated genotype/phenotype/experimental information is stored in the European Genome-phenome Archive (EGA) under controlled access with Dataset ID EGAS00001005830.

### 2.4 RNA-sequencing primary data analysis

Adapter sequences and low-quality base calls were removed from fastq files using cutadapt 3.4 via TrimGalore 0.6.6 at no other default parameters than the --paired flag [28]. Trimmed fastq files were mapped to the human genome version hg38.95 using STAR 2.7.0 at default parameters and --outSAMtype BAM SortedByCoordinate [29]. After indexing using samtools [30] at default parameters, PCR-duplicates were removed from the bam files using umi-tools dedup with the flags --spliced-is-unique, --paired and --output-stats (Supplementary Table 1)[31]. Strandedness was verified via RSeQC’s infer_experiment [32]. After indexing the deduplicated bam files, reads were counted for overlap with hg38.95 genes via HTSeq with parameters --format bam --order pos and --stranded=yes [33]. EPIC, quanTIseq and xCell algorithms were applied to the count tables to verify that the cell type compositions were similar at the two time points [34–36]. GATK HaplotypeCaller and Picard GenotypeConcordance were used to check correct matching of samples from the same patient [37,38].

Splice analysis was performed using rMATS v4.1.0 [39] via the same gtf as for STAR/HTSeq with the parameters and flags: -t paired --readLength 150 --variable-read-length --novelSS -- libType fr-firststrand --statoff.

### 2.5 Comparative DM1 datasets

To validate our findings with previously published datasets, gene counts were extracted from the associated publications [11,12] or were obtained from count tables or raw data as described above from GEO entries GSE85984 and GSE67812 [15,25]. The dataset EV10 from Signorelli et al was obtained for the association between gene expression (logFC) and body-or performance test in Duchenne Muscular Dystrophy patients[22].

To compare DM1 samples to controls a two-sided two-sample Wilcoxon test was performed using *row_wilcoxon_twosample* in matrixTests R-package on normalized, log-transformed gene counts [40].

### 2.6 Statistical analysis

All statistical analyses were carried out in R [41]. For gene expression analysis, firstly genes with low read counts before and after CBT were filtered using edgeR filterByExpr with group = before/after CBT and min.count = 50 [42]. Following, normalized logCPM values and weights were calculated from the filtered read counts with Voom in Limma [43].

We first set out to explore differential gene expression before and after the CBT intervention. Gene expression values from Voom (in logCPM) were separately modelled using mixed effect models with before/after CBT (categorical) as fixed effect and patient identity as random effect [1]. Gene weights were also carried over from Voom. This analysis has been implemented using *lme* in the lme4-wrapper lmerTest [44,45]. lmerTest estimates a p-value for the contribution of fixed effects to the model via the Satterthwaite’s degrees of freedom method. Fit parameters were extracted with R base *summary* and p-values were FDR corrected via the Benjamini & Hochberg method with stats *p.adjust* [46].

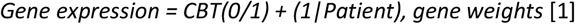

In order to study the cohort heterogeneity of gene expression changes, we calculated the logCPM based difference in gene expression between before and after the intervention for each patient for the 560 genes significantly associated with the CBT predictor of [1] (adjusted-p < 0.05). Patients and genes with similar expression changed were clustered using the R package heatmap3 based on the complete linkage method for hierarchical clustering, with gene expression values being centered and scaled per gene [47].

Next, using the same methodology as for the CBT intervention effect, we set out to explore the associations of the different clinical outcome measures and CTG-repeat length with gene expression. For this purpose, we separately modelled gene expression values with either one of the outcome measures or the CTG-repeat length (at trial start, [2]) as fixed effect and patient identity as random effect. The categorical CBT covariate (before/after) was included for each fit to correct for differences between the two time points.

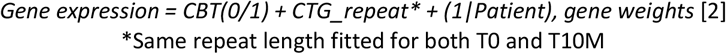

To achieve an overarching measure for CBT response, first the changes for each outcome measure were calculated per patient by subtracting the value after 10 months CBT from that at baseline. Where applicable, outcome measures were multiplied by -1 in order to always associate positive changes with an improved health status. Using R base *scale* [41], the changes per outcome measure were then scaled without centering to account for the different scales of the outcome measures. Finally, for each patient a ‘Compound Response’ score was calculated based on the mean of all scaled outcome measures. L5ENMO, the mean activity during rest, was a control parameter in OPTIMISTIC and was excluded from this analysis.

Analogous to the methodology described for the outcome measure association analyses, genes associated with overarching clinical response were identified by fitting separate mixed effect models for each gene with the two fixed effects CBT (categorical) and Compound Response, as well as patient identity as random effect [3]. Notably, the Compound Response variable has only been fitted for gene expression after the CBT intervention (zero at baseline). As such, the Compound Response predictor reflects the difference between the two time points that can be attributed towards therapy responsiveness, while accounting for non-therapy specific differences by including the categorical CBT predictor.

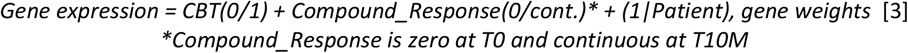

For the splicing analysis, the PSI values for splice exclusion (SE) events were extracted from the rMATS output and fitted in linear models similar to those for gene expression. Splice events were filtered by excluding exons from the analysis with less than three mapping reads and one junction spanning read in at least 14 samples.

The R package ggplot2 was used for representation in volcano- and scatter plots [48]. The R package VennDiagram was used to generate the Venn diagram in Figure 5 [49].

### 2.7 Gene-set enrichment analysis

Gene-set enrichment analyses have been independently implemented for the gene-sets associated with CBT, CTG-repeat length, Compound Response scores and the genes significantly associated with both CTG-repeat length and Compound Response using gProfiler [50]. For the CBT, CTG-repeat length and Compound Response associated genes, the 500 genes with the lowest nominal p-values were ordered (decreasing) based on their absolute regression coefficients. Subsequent enrichment analyses were implemented using the R client of gProfiler with the parameter orderd_querey = TRUE against a custom background of 10,292 genes expressed in our samples. Multiple testing correction was based on the default setting ‘g_SCS’. We tested for enrichment (one-sided) pathways within the WikiPathway database. The gene-set associated with both CTG-repeat length and Compound Response was based on an FDR threshold of 10% for the respective regression coefficients, resulting in a gene set of 311 genes. A regular, non-order weighted ORA analysis was run for this gene-set with ordered_quere = FALSE. Similar to the other analyses, one-sided (enrichment) pathway discovery was based on the WikiPathway database with the default setting ‘g_SCS’ to correct for multiple testing. For all enrichment analyses only significant pathways (p-adjusted < 0.05) are reported.

Exact scripts and the resulting datasets of the statistical analyses are available via https://github.com/Remcovc/DM1_blood_RNAseq/

## 3. Results

### 3.1 Sample selection, procedure and analysis of outcome measures

For the identification of blood biomarkers that are associated with the clinical response to the CBT intervention, we selected 27 patients from the OPTIMISTIC cohort. These patients reflected a uniform continuous distribution of therapy responses as assessed by the primary trial outcome, the DM1-Activ-c questionnaire.

The selected set consisted of 14 females and 13 males aged 19-63 years and represented a range of CTG-repeat lengths (Supplemental Figure S1). All selected patients received CBT, and mRNA-sequencing profiles were obtained at baseline and after 10 months of CBT, the primary endpoint of the OPTIMISTIC trial. These patients featured variable extents of improvement/deterioration after CBT based on the different outcome measures that were collected during the trial (Figure 1).

**Figure 1:**
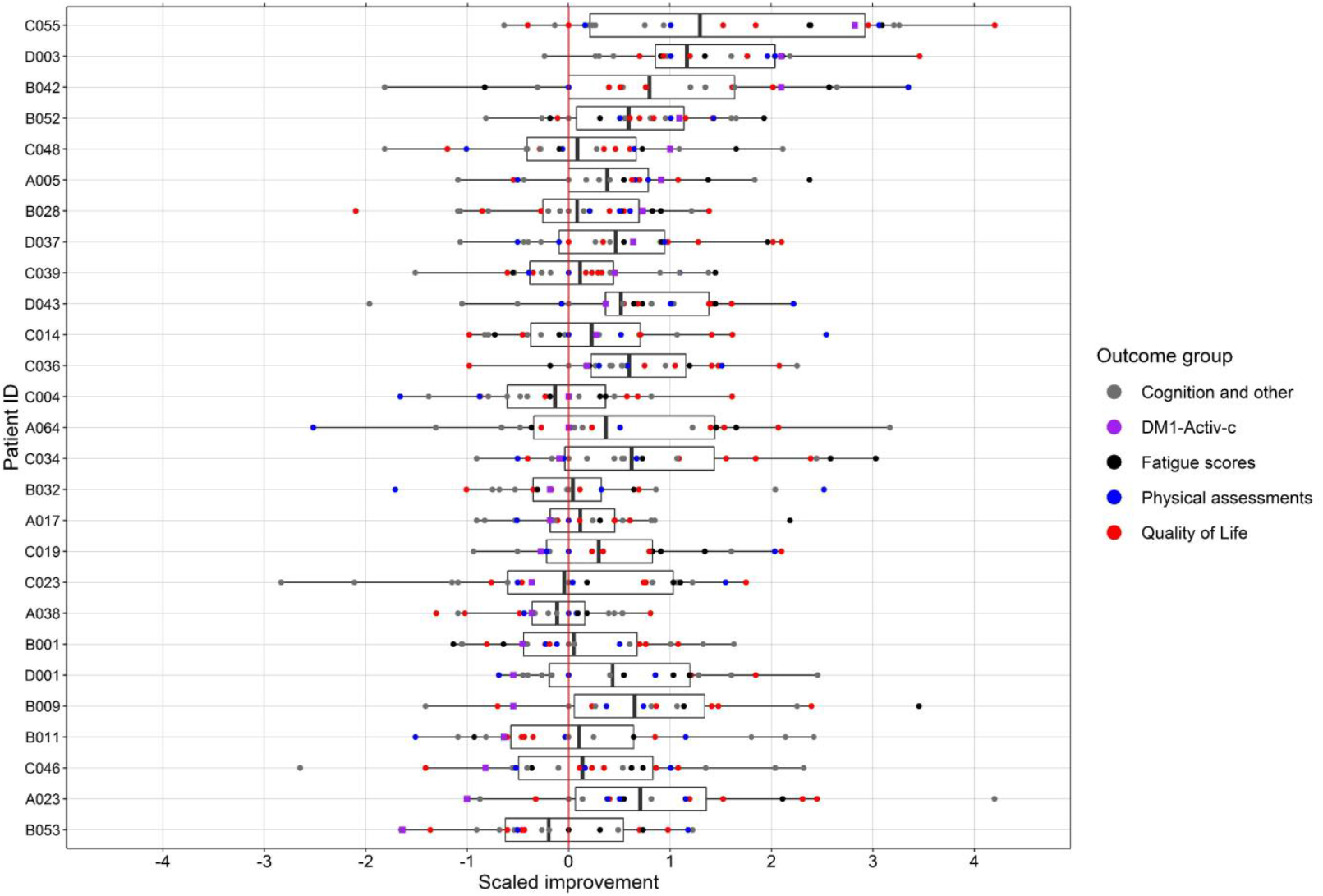
Distribution of changes in outcome measures per patient. Per outcome measure, changes between baseline and 10 months of CBT were scaled by the root mean square. Additionally, for some outcome measures a sign-adjustment was performed so that an increased score is always associated with improved health status. All outcome measures are shown per patient, where patients were ordered along the y-axis by their change in DM1-Activ-c scores (purple squares). Boxes enclose the 25^th^ to 75^th^ percentiles, divided by a thick line that represents the mean compound response score. The whiskers represent the lowest/highest value no further than 1.5 times the interquartile range. **Quality of life (red):** Myotonic Dystrophy Health Index, Individualized Neuromuscular Quality of Life Questionnaire, Adult Social Behavioural Questionnaire, Illness Management Questionnaire, Checklist individual strength – Subscale activity. **Physical assessments (blue):** Six Minute Walk Test, BORG Scale, Accelometery measures **Fatigue scores (black):** Fatigue and Daytime Sleepiness Scale, Checklist Individual Strength – Subscale fatigue, Jacobsen Fatigue Catastrophizing Scale **Cognition and other (gray):** Trail Making Test, Stroop Color-Word Interference Test, McGill Pain Questionnaire, Beck Depression Inventory – Fast Screen, Social Support – Discrepancies and Negative Interactions, Apathy Evaluation Scale – Clinical Version, Self-Efficacy Scale 28

### 3.2 Marked changes in gene expression after CBT

We first studied the molecular changes that occurred after the CBT intervention by comparing the mRNA expression levels in blood before and after the CBT intervention. We found that 560 genes were significantly up- or downregulated after CBT (277 genes down, 283 up, Figure 2A). Hierarchical clustering of patients based on the changes in these 560 genes revealed substantial molecular heterogeneity within this 10 months timeframe, which was not in concordance with changes in clinical phenotypes (Figure 2B). The four most significant genes were *GGCX, ZNF16, SERBP1 and SLC39A8* (Figure 2C). Biological pathways significantly associated with these genes were limited to an immunological pathway and an electron transport chain in mitochondria (Table 1).

**Table 1:** Gene set enrichment analysis.

| query | Adjusted p-value | Term size | Intersection size | Term ID | Term Name | Intersection with WikiPathway |
| --- | --- | --- | --- | --- | --- | --- |
| CBT_ORA | 0.014 | 49 | 5 | WP:WP5039 | SARS-CoV-2 Innate Immunity Evasion and Cell-specific immune response | *CXCL1, PPBP, PF4, CXCR2, CXCL5* |
| CBT_ORA | 0.037 | 91 | 8 | WP:WP111 | Electron Transport Chain (OXPHOS system in mitochondria) | *MT-ND3, ATP5F1E, MT-ND1, MT-ATP6, MT-CO2, MT-CO3, MT-ND4, MT-CO1* |
| CTG_ORA | <0.001 | 46 | 10 | WP:WP286 | IL-3 Signaling Pathway | *FOS, CSF2RB, GAB2, HCK, LYN, MAPK3, RAF1, STAT3, STAT5B, PIK3CD* |
| CTG_ORA | 0.001 | 49 | 10 | WP:WP395 | IL-4 Signaling Pathway | *FOS, IRS2, CEBPB, GAB2, NFIL3, MAPK3, STAT6, STAT3, STAT5B, PIK3CD* |
| CTG_ORA | 0.002 | 26 | 3 | WP:WP4564 | Neural Crest Cell Migration during Development | *MMP9, EPHB4, FOS* |
| CTG_ORA | 0.003 | 28 | 3 | WP:WP4565 | Neural Crest Cell Migration in Cancer | *MMP9, EPHB4, FOS* |
| CTG_ORA | 0.004 | 29 | 7 | WP:WP3972 | PDGFR-beta pathway | *FOS, MAPK3, RAF1, MAP2K4, STAT6, STAT3, STAT5B* |
| CTG_ORA | 0.006 | 29 | 2 | WP:WP4808 | Endochondral Ossification with Skeletal Dysplasias | *MMP9, ALPL* |
| CTG_ORA | 0.006 | 29 | 2 | WP:WP474 | Endochondral Ossification | *MMP9, ALPL* |
| CTG_ORA | 0.006 | 56 | 2 | WP:WP2324 | AGE/RAGE pathway | *MMP9, ALPL* |
| CTG_ORA | 0.008 | 48 | 10 | WP:WP304 | Kit receptor signaling pathway | *FOS, GAB2, SH2B2, LYN, MAPK3, RAF1, STAT3, STAT5B, CRK, RPS6KA1* |
| CTG_ORA | 0.011 | 37 | 8 | WP:WP127 | IL-5 Signaling Pathway | *FOS, CSF2RB, LYN, MAPK3, RAF1, STAT3, STAT5B, RPS6KA1* |
| CTG_ORA | 0.013 | 31 | 7 | WP:WP313 | Signaling of Hepatocyte Growth Factor Receptor | *FOS, PTEN, MAPK3, RAF1, STAT3, CRK, PXN* |
| CTG_ORA | 0.015 | 110 | 16 | WP:WP3929 | Chemokine signaling pathway | *CXCR2, TIAM2, HCK, NCF1, LYN, ARRB2, GNB2, PREX1, MAPK3, RAF1, STAT3, STAT5B, CRK, WAS, PXN, PIK3CD* |
| CTG_ORA | 0.031 | 15 | 3 | WP:WP3599 | Transcription factor regulation in adipogenesis | *IRS2, CEBPD, CEBPB* |
| CTG_CR_ORA | 0.002 | 110 | 13 | WP:WP3929 | Chemokine signaling pathway | *WAS, PXN, HCK, MAPK3, PREX1, ARRB2, PIK3R5, VAV1, NCF1, CXCL16, PIK3CD, GNB2, GRB2* |
| CTG_CR_ORA | 0.006 | 46 | 8 | WP:WP286 | IL-3 Signaling Pathway | *GAB2, CSF2RB, HCK, MAPK3, PTPN6, VAV1, PIK3CD, GRB2* |
| CTG_CR_ORA | 0.009 | 48 | 8 | WP:WP304 | Kit receptor signaling pathway | *GAB2, MAPK3, PTPN6, RPS6KA1, VAV1, SH2B2, JUNB, GRB2* |
| CTG_CR_ORA | 0.016 | 115 | 12 | WP:WP306 | Focal Adhesion | *ACTN1, PXN, HCK, MAPK3, CCND2, VASP, TLN1, VAV1, ZYX, ITGA5, PIK3CD, GRB2* |
| CTG_CR_ORA | 0.032 | 31 | 6 | WP:WP3937 | Microglia Pathogen Phagocytosis Pathway | *HCK, PTPN6, VAV1, NCF1, ITGB2, PIK3CD* |

**Figure 2:**
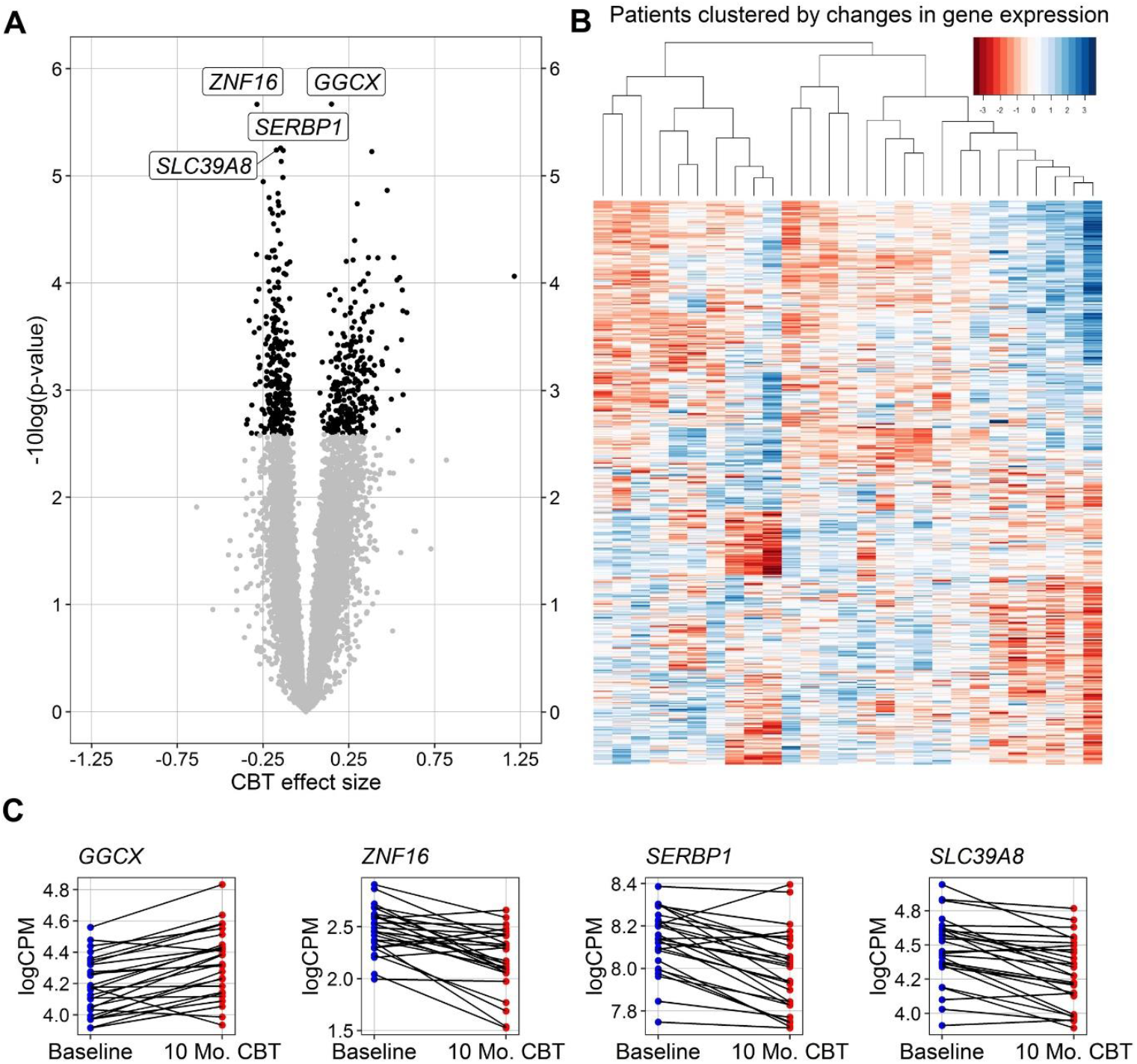
Changes in gene expression after cognitive behavioural therapy. A linear mixed effect model was fitted for each gene, estimating the fixed effect of CBT while accounting for (random) effects of the individual. The p-values for the fixed effect were estimated via Satterthwaite’s degrees of freedom method and FDR-corrected. A) Volcano plot of significance (-log10 of the nominal p-value) and the effect size for changed expression after 10 months of CBT. Genes for which the effect size of CBT is significant (FDR < 0.05) are visualized in black. B) Heatmap of changes in normalized logCPM values between the baseline and the 10 months assessment for the 560 genes significantly associated with the CBT effect size. Patients and genes were clustered based on the complete linkage method for hierarchical clustering, values were centered and scaled per gene. C) Expression values (logCPM) at baseline(blue) and after CBT(red) of the four genes with the lowest nominal p-values from panel A.

### 3.3 CTG-repeat length associations reflect molecular dysregulation in blood across different studies

Having a readily accessible fingerprint reflecting the molecular dysregulations associated with DM1 are potentially of high value for both clinical and research settings. To this end, we studied the correlations of blood-based gene expression with the disease causing CTG-repeat length assessed at the start of the trial. The assessments were based on small pool PCR, *Acil* digestion and Southern blot, as opposed to (estimations) of CTG repeat length at birth or disease onset. This was done to minimize potential confounding effects related to the progressive nature of the disease and to assure homogeneity in measurement methodology.

Based on this approach, we identified 608 genes significantly associated with the CTG-repeat at an FDR cutoff of 5% (474 positively, 134 negatively, figure 3A). The four most significant genes *RNF170, IRS2, NDE1* and *PRIMPOL* showed a clear linear relationship between the CTG-repeat length at baseline and expression values, both before and following the intervention (Figure 3B). Most enriched pathways were related to immunological processes (IL-3, IL-4, IL-5 signaling; chemokine signaling pathway), yet also pathways related to adipogenesis, hepatocyte signaling and AGE/RAGE were discovered. Interestingly, the gene *MMP9* was among several of the CTG-repeat associated pathways.

**Figure 3:**
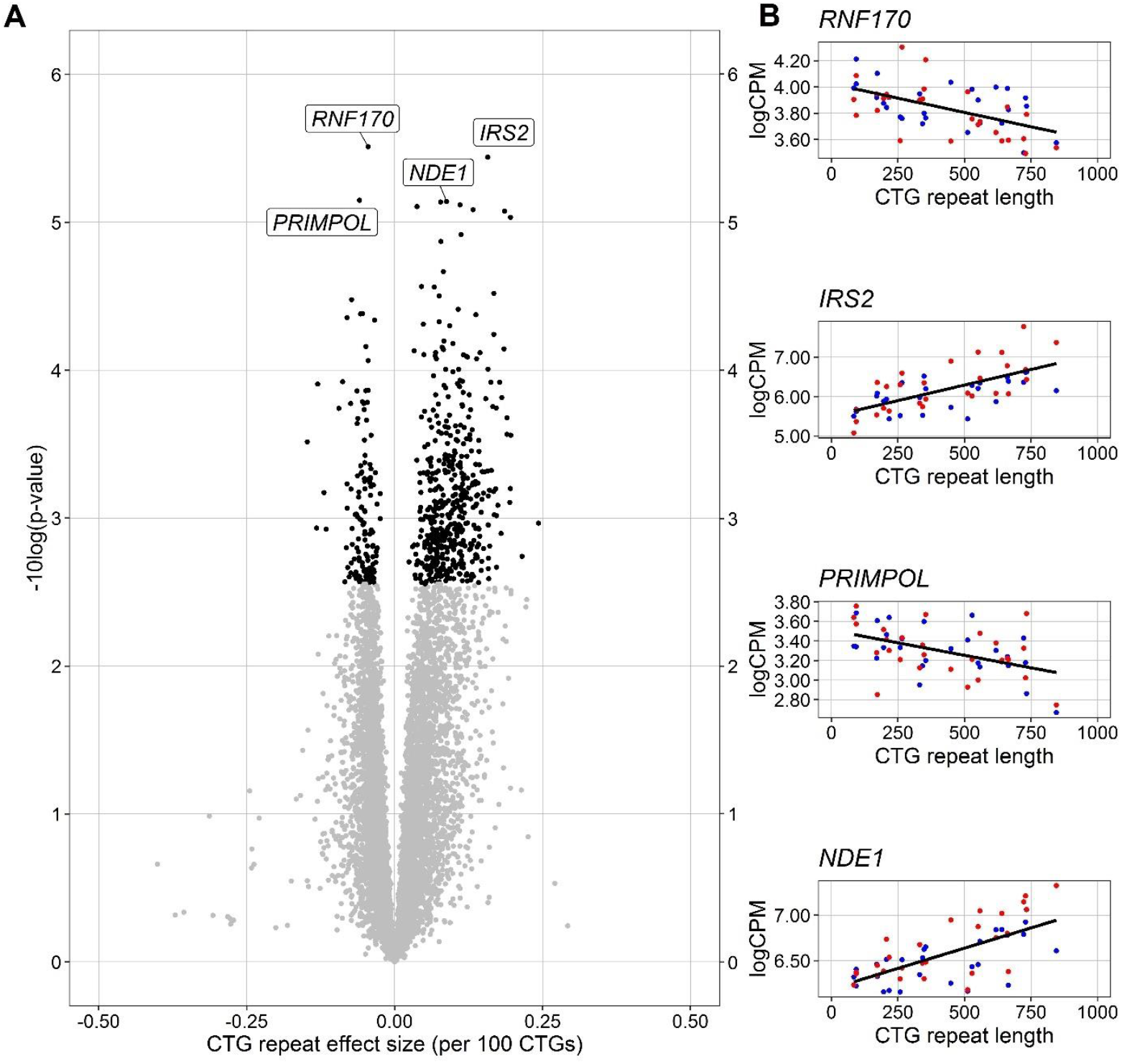
Gene expression levels associated with CTG-repeat length. For each gene a mixed effect model was fitted with before/after CBT and CTG-repeat length as fixed effects, while accounting for (random) effects of the individual. The p-values for the fixed effects were estimated via Satterthwaite’s freedom method and FDR corrected. A) Volcano plot of significance (-log10 of the nominal p-value) and effect size of the CTG-repeat length (per 100 CTGs) on gene expression. Genes for which the effect of CTG-repeat is significant (FDR < 0.05) are visualized in black. B) For the four genes with the lowest nominal p-values from panel A, the gene expression values (logCPM) are plotted against the CTG-repeat length. Blue dots represent baseline expression values, red dots expression values after CBT. The regression line is fitted over all values, independent of the timepoint.

To validate that these findings are disease relevant, we compared the CTG-repeat length effect size with effect sizes reflecting the differences in gene expression between DM1 and controls in various published datasets (Supplemental Figure S3). We found a strong correlation with a study that performed mRNA sequencing on DM1 and control blood-samples (Pearson Rho: 0.59, figure S3A [25]). However, similar correlations were not observed for studies profiling DM1 and control tissues other than blood (heart fig S3B [15], brain fig S3C [12], muscle fig S3D [11]). Neither were correlations observed with inferred *MBNL* activity or muscle strength measures from another study (fig S3E and S3F [11]). Interestingly, the CTG-repeat length effect size did correlate well (Pearson Rho: 0.42, Figure S3H) with the effect size of body measurements DMD blood samples, while not correlating with physical test assessments (fig S3G)[22]. In the latter study, comparisons with physical tests and body measures were independently based on the first principal component of a set of different clinical assessments of DMD patients. Thus, while DM1-related molecular dysregulations in blood can be validated in other studies, even from other neuromuscular disorders, they do not necessarily reflect expression dysregulation observed in other tissues.

Since DM1 is known as a splicing disease, we also studied the association of the CTG-repeat length with alternative splicing events. Here, four events in three genes (*RBM39, FLNA* and *CTSZ*) reached an FDR threshold of <5%, (Supplemental Figure S4). Given the limited and small effects observed we did not further explore these associations.

### 3.4 Non-significant associations between gene expression and individual clinical outcome measures

Next we studied phenotype-genotype relationships by estimating the associations of gene expression values with individual clinical outcome measures used in the OPTIMISTIC trial. Here we found virtually no significant associations between gene expression and clinical outcomes. Shown in Supplemental Figure S2 are the associations of gene expression values with the DM1-Activ-c questionnaire. The four genes with the lowest nominal p-values were *SREBF2, ZNF283, SF3B3* and *GSKIP*.

### 3.5 Significant associations with average clinical response

To account for the heterogenic changes in individual outcome measures, we calculated a compound CBT response score that reflects the average therapy response of all outcome measures used in OPTIMISTIC (Figure 1). The compound response score can be interpreted as an estimate of an overall difference in capacity between the end and the start of the intervention. We were able to identify 1176 genes significantly (FDR < 0.05) associated with the compound response score (384 positive, 792 negative, Figure 4A). The four most significant hits (*PPP1R9B, CSNK1G2, PPP6R1, FBXO48*) show a clear linear relationship between changes in gene expression during the trial and the Compound Response score (Figure 4B). No enriched pathways were identified for this gene set.

**Figure 4:**
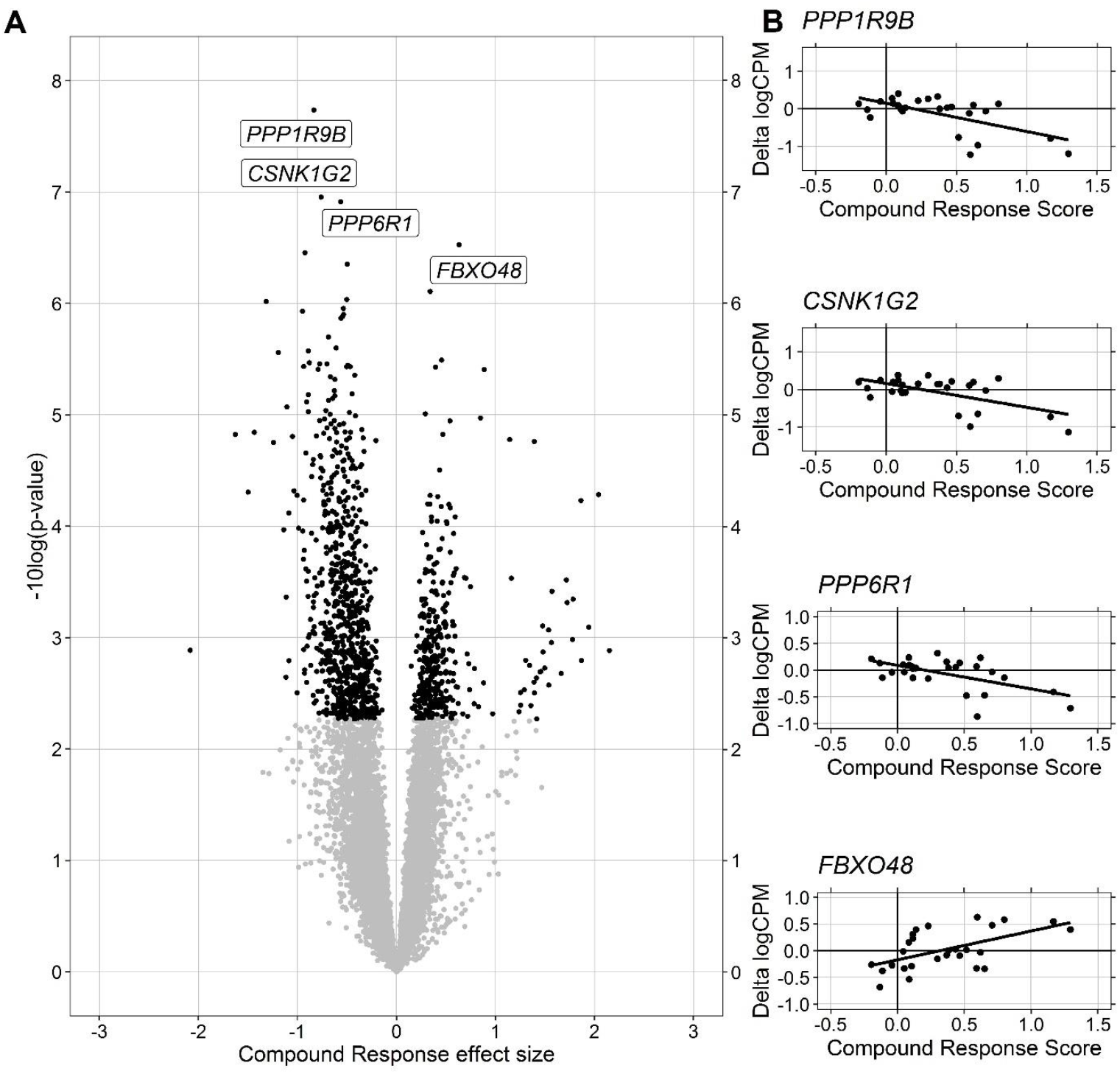
Gene expression association with Compound Response scores. For each gene, a mixed effect model was fitted with before/after CBT and Compound Response Scores as fixed effects, while accounting for (random) effects of the individual. Compound Response Scores were fitted for gene expression values after CBT and set to be zero at baseline, the effect size of this covariate therefore expresses changes in gene expression compared to the baseline values that are associated with clinical response. The p-values for the fixed effects were estimates via Satterthwaite’s freedom method and FDR corrected. A) Volcano plot of significance (-log10 of the nominal p-value) and the effect size of the Compound Response Score on gene expression. Genes for which the effect size of Compound Response is significant (FDR < 0.05) are visualized in black. B) For the four genes with the lowest nominal p-values from panel A, the changes in gene expression (delta logCPM after-before CBT) are plotted against the Compound Response scores.

### 3.6 Clinical improvement linked to reversal of disease-induced gene expression

Since we were able to identify genes significantly associated with both the CTG-repeat length as well as the average clinical response, we were interested in their intersection. Among the significant hits of both analyses, we found an overlap of 97 genes (Figure 5A). To further investigate this relationship, we plotted the effect size of the Compound Response score against the effect size of the CTG repeat length for each expressed gene (Figure 5B). For the 97 genes significantly associated with both predictors a remarkable pattern emerged: genes that were lower expressed in patients with a long CTG repeat showed an increase in expression levels in the patients with a good clinical response and vice versa. This anticorrelation pattern was confirmed by analyzing an earlier dataset comparing gene expression in DM1 and control blood samples [25]. Here the 97 genes showed a similar association with the DM1 phenotype as has been found with the CTG-length in our study, confirming that the gene expression of patients with a good CBT response changed into the direction of the levels observed in healthy controls (Figure 5C). This remarkable relationship could not be explained by possible confounding between CTG-repeat length and Compound Response, as the Compound Response effect size is only slightly affected by first regressing out the CTG-repeat length effect (Supplemental Figure S5). The four genes most significantly associated with both the CTG-repeat length as well as the Compound Response Score (either FDR < 0.025) were *DNAJB12, HDAC5, TRIM8* and *ZNF22*. Enriched biological pathways were mostly related to immunological pathways (Chemokine and IL-3 signaling, Microglia Pathogen Phagocytosis Pathway; table 1).

**Figure 5:**
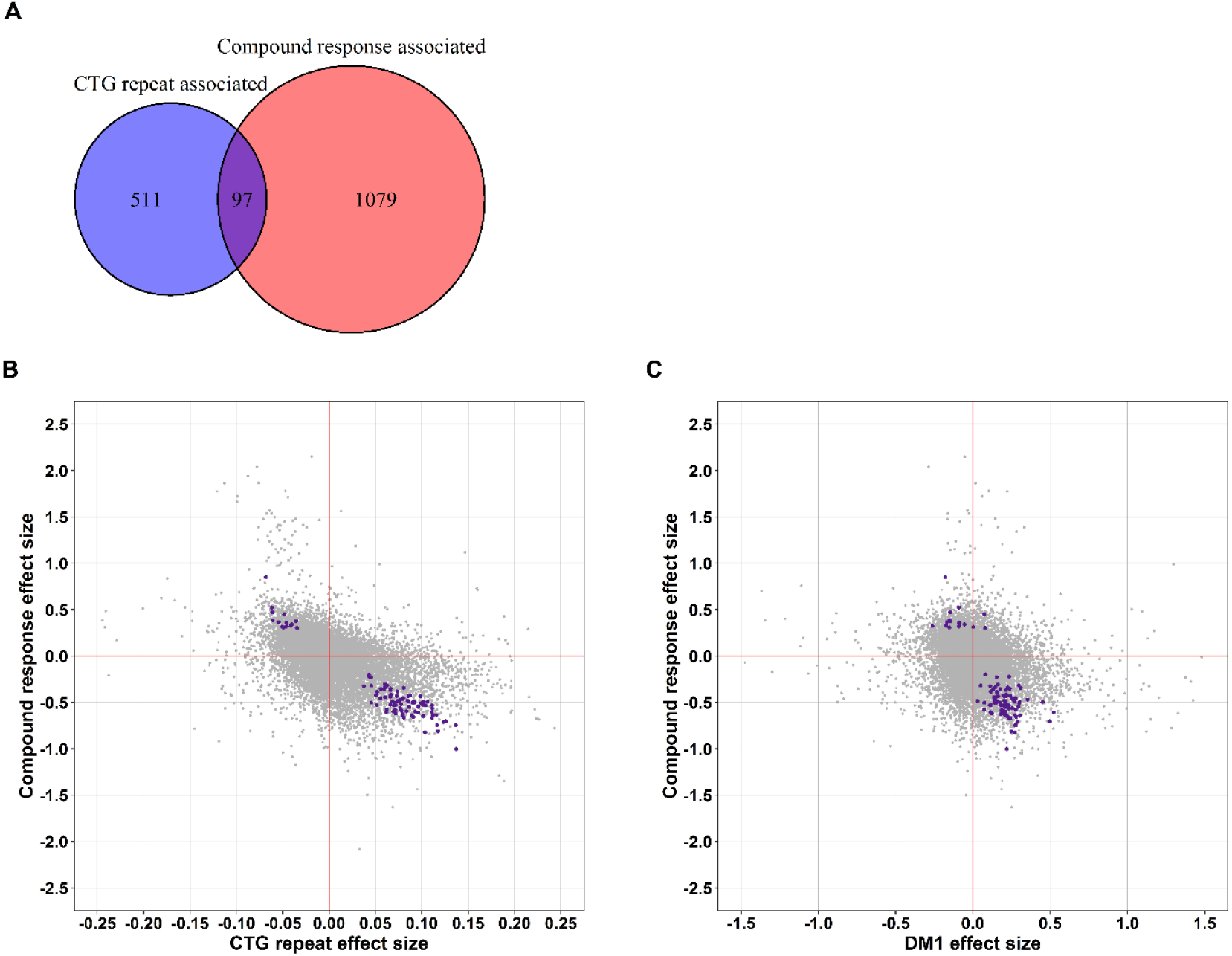
Clinical improvement is linked to normalization of expression of CTG-repeat associated genes. Linear mixed effect models were fitted for each expressed gene, with CBT as fixed effect, patient as random effect, and either CTG repeat or Compound Response as predictor. P-values for the regression coefficients were estimated via Satterthwaite’s degrees of freedom and considered significant for values smaller than 0.05 after FDR correction. Furthermore, differences in gene expression of blood samples from DM1 patients and controls were calculated based on an external study using a Wilcoxon signed-rank test on normalized, log-transformed gene counts. A) Venn- diagram illustrating the number of significant genes associated with CTG repeat length and Compound Response, as well as their overlap (disease relevant changes). B) For all expressed genes, the regression coefficients of the Compound Response scores are plotted against the regression coefficients of the CTG repeat lengths. For illustrative purposes the regression coefficients of the CTG repeat have been multiplied by 100. Furthermore, the x-axis has been scaled between -0.25 and 0.25, removing 12 outliers from the figure. Colored in purple are the genes for which both regression coefficients were significant (FDR < 0.05). C) For all expressed genes, the Compound Response effect size is plotted against the DM1 effect size based on an external study comparing blood expression profiles from DM1 patients and controls [25]. For illustrative purposes the x-axis has been scaled between -1.5 and 1.5, removing 6 outliers from the figure.

In summary, these results suggest that for a subset of genes significantly associated with the biochemical phenotype caused by the CTG-repeat expansion a reversal of disease-induced gene expression occurred in clinical responders. The association with both molecular dysregulation and clinical response makes this subset of genes highly relevant for the discovery of novel therapeutic targets.

## 4. Discussion

The purpose of this study was the identification of DM1 specific therapeutic biomarkers in peripheral blood. The multisystem nature of DM1 is known to be reflected by laboratory abnormalities of peripheral blood, making it together with its accessibility a promising tissue for biomarker studies in this disease [26]. Hence, we used blood samples of 27 carefully selected DM1 patients from the OPTIMISTIC cohort to study the associations of gene expression with disease severity as well as the response towards the CBT intervention. The careful selection was done to facilitate generalizability of the study findings, as well as to be able to establish linear associations with therapy response. We identified substantial heterogeneity in molecular expression profile changes after the 10 month CBT-intervention. Furthermore, we identified gene sets that were significantly associated with the disease causing CTG-repeat as well as with the average response towards the CBT intervention across different clinical outcome measures. Most interestingly, an overlap of 97 genes among these latter two gene sets has been identified, showing a clear trend of more normal expression levels in clinical responders. Based on these different gene sets several biological pathways relevant to DM1 have been discovered, as well as specific genes/gene-families with ties to neuro(-muscular) disorders.

The OPTIMISTIC study has shown that DM1 patients significantly improve in their (capacity) for activity and social participation after the CBT intervention [18]. It was furthermore hypothesized that CBT may directly or indirectly improve other biological systems affected by the disease. This hypothesis has been confirmed for muscles of the lower extremity, showing an increase in cross-sectional area as a result of the intervention [51]. Here we set out to further explore this hypothesis by investigating changes on the molecular level. These changes may be the result of increased physical activity, which has been linked to differences in gene expression in previous studies [52], but may also be a more direct effect of the psychotherapeutic CBT intervention [53].

In line with the results of an earlier study, we have illustrated that the clinical response towards the CBT intervention was rather heterogenous [21]. A novel addition to this finding was the illustration that this heterogeneity extends towards changes in molecular profiles within a 10 month timeframe. Importantly, this heterogeneity could not be explained by changes in cellular composition of the blood samples between the two time points, as similarity of cell type composition has been verified. Additionally, this heterogeneity could not be explained by changes of different outcome measures such as the DM1-Activ-c scale. While the CBT intervention likely played a part in this heterogeneity, the magnitude of this contribution could not be assessed due to the lack of a control group. Other factors, such as ageing or seasonal effects may also have contributed to this finding.

Across the different gene sets identified in this study several of the most significant genes (*SLC39A8, IRS2, FBXO48)* and one WikiPathway (Transcription factor regulation in adipogenesis) were associated with insulin signaling or more broadly related to metabolism/adipogenesis [54–56]. Dysregulation of insulin signaling has been associated with clinical features of DM1 and is an actively ongoing field of investigation [57]. Aberrant insulin signaling has also been found in other diseases of the nervous system such as depression, with indirect improvements being observed after CBT [58]. Interestingly, the anti-diabetic drug Metformin has been shown to improve the mobility of DM1 patients with effect sizes of the six minute walk test comparable to those observed in the OPTIMISTIC study [59]. With increasing therapeutic interest in this area, our findings suggest that disease relevant insulin signaling can be studied on a molecular level in blood samples, highlighting the utility of peripheral blood in this setting.

Similarly, across most of the gene sets we identified several WikiPathways associated with immunological functions (cell-specific immune response, chemokine signaling pathway, IL-3, 4 and 5 signaling). Whilst this may be in part due to a bias introduced by the profiled tissue, the immune system likely plays a role in the DM1 pathophysiology like for many other chronic diseases [60]. As such, blood sample-based immunology studies may be an interesting field of future investigation.

The intersection of the genes significantly associated with the disease causing CTG-repeat as well as the average CBT response across different outcome measures, revealed a subset of 97 genes. These genes are of particular interest for the identification of therapeutic biomarkers, as they are not only associated with the DM1 phenotype, but also showed normalization of expression levels in clinical responders. Among the genes most significantly associated with both were *HDAC5, DNAJB12* and *TRIM8*. In total the subset of 97 genes consisted of two HDACs (histone deacetylases, *HDAC5, HDAC7*). HDACs play an important role in transcriptional regulation and compounds that inhibit HDAC enzymes are being studied for their potential effect on a range of human diseases, including neurological disorders [61]. The DNAJB12 protein is member of the Heat shock protein family, with some evidence supporting positive effects of their induction for muscular dystrophy and other muscle wasting conditions [62]. The TRIM family protein TRIM72 has been shown to be an essential component of cellular membrane repair in muscles, with evidence supporting some positive effects in mouse models of muscular dystrophy [63]. Authors of the same study suggest the potential of other TRIM family members as potential targets in similar disease states, which may support the further investigation of *TRIM8* in DM1. Although mostly associated with therapy response, *RARA* and *RXRA* were also among the overlapping 97 genes. Stimulating retinoic acid signaling has been linked to muscle regeneration in mouse models via increased proliferation of fibro/adipogenic progenitor cells, highlighting the relevance of this pathway as another potentially DM1 relevant drug target [64]. Taken together, these findings confirm the value of whole blood-based expression profiling for the discovery of therapeutic biomarkers in DM1.

Interestingly, the genes significantly associated with the CTG-repeat showed a moderate correlation with the genes associated with DMD body measurements of an external study. We hypothesize that these body measures are likely also correlated with age, which in turn reflects disease progression. This suggests that some of our results may therefore not be DM1 specific, but rather reflect non-specific molecular dysregulations shared across different (neuromuscular) disorders. This hypothesis is supported by the significant association between the CTG-repeat with *MMP9*, which is known to be a non-specific biomarker that has for instance been linked to cardiac remodeling after myocardial infarction, inflammation and DMD [65,66]. We therefore deem further exploration of shared dysregulations as highly valuable, as this may lead to the discovery of therapeutic targets relevant to a variety of diseases.

Although DM1 is known as alternative splicing disease, only four splice events have been significantly linked to the disease causing CTG-repeat in this study. This may be the result of low *DMPK* expression in blood, which we also believe to be an important contributing factor towards the finding that our CTG-repeat effect size did not correlate with DM1 effect sizes of other tissues from external studies. So, while whole-blood based transcription profiling can identify disease relevant molecular dysregulations, these dysregulations do likely not fully reflect the dysregulations observed in other tissue types. Yet, we found a high correlation of the CTG-repeat effect size with the DM1 phenotype effect size of a different blood based study, as well as with a principal component derived body measure association of a Duchenne Muscular Dystrophy blood based study. While the former validates our findings, the latter suggests the possibility of shared, disease relevant, dysregulations across different neuro(muscular) disorders detectable in peripheral blood. If true, associated pathways might reveal highly interesting targets for drug discovery, as they may have a positive influence on multiple diseases.

### 4.2 Limitations of this study

To find disease relevant gene-expression in blood, we searched for linear associations with the disease causing CTG-repeat length. While the CTG-repeat length is thought to be the main driver of molecular dysregulation, associations between the progenitor allele length of the CTG repeat with several clinical outcome measures, including DM1-Activ-c and six-minute walk test, have been found to be only weak-moderate [13]. In line with the previously published challenges to directly relate gene expression to clinical phenotypes, we were not able to find significant, direct associations between clinical outcome measures and gene expression. Still, among the most significant genes that were associated with the DM1-Activ-c questionnaire was *GSKIP*, a gene encoding for an inhibitor protein of the known DM1 drug target GSK3-β [67–69]. Given the biological relevance of this finding we deem it likely that the current study design was underpowered to study the association of gene expression with individual clinical outcome measures, especially when taking clinical and molecular heterogeneity into account.

The clinical heterogeneity in therapy response may in part be explained by the personalized nature of the CBT intervention, with therapy foci being tailored towards the needs and wishes of the individual patient. As a consequence, one might expect different outcome measures to be more appropriate for CBT efficacy assessments for different patients. Yet, the identification of molecular signatures associated with therapy response necessitates the use of the same clinical outcome measure. For this reason, and to average out some of the uncertainty inherently associated with the recording of the different outcome measures, we settled on the use of the compound response score. However, we acknowledge that this combined score is biased by the outcome measures that were used in OPTIMISTIC.

Even though we statistically corrected for non-specific molecular changes between the two time points, the lack of RNA-seq profiles from the OPTIMSITIC control arm makes it difficult to state with certainty that the observed molecular changes are due to the therapy itself. However, this does not discount their value as potential therapeutic targets, as they are, regardless of the mediation factor, significantly associated with improved clinical status. Moreover, for this reason we deemed studying the RNA-seq profiles of the OPTIMISTIC control arm to be less valuable, as these patients did not significantly clinically declined over the 10 month time frame [18].

### 4.3 Conclusion

Starting from DM1 specific disease determinants, the OPTIMISTIC study has shown that patient tailored CBT can increase the health status of DM1 patients by improving social participation and activity. It was furthermore hypothesized that the CBT intervention positively challenges the biological system, which has already been confirmed by increased cross-sectional area for muscles of the lower extremities. Making use of the clinical heterogeneity in therapy response, we here additionally confirmed disease relevant molecular changes in peripheral blood. Not only do our results highlight the utility of peripheral blood to study the multisystem nature of the disease, but also generated the foundation for an upcoming, multi-omics based drug repurposing study.

## Data Availability

All raw sequencing data and associated genotype/phenotype/experimental information is stored in the European Genome-phenome Archive (EGA) under controlled access with Dataset ID EGAS00001005830

https://ega-archive.org/studies/EGAS00001005830

## 5. Acknowledgements

We are grateful to the Newcastle MRC Centre for Rare & Neuromuscular Diseases Biobank, and in particular to dr. Dan Cox, for assistance in the storage and delivery of the patient samples used in this study. We also like to thank Dr. Rick Wansink and Ing. Walther van den Broek for helpful discussions and their work to isolate RNA. Furthermore, we would like to thank dr. Pietro Spitali for the helpful discussions regarding the shared dysregulations observed with Duchenne Muscular Dystrophy.

## 7. Author contributions

**Remco van Cruchten:** Data Curation, Methodology, Validation, Formal analysis, Writing – Original Draft, Visualization;

**Daniël van As:** Methodology, Validation, Formal analysis, Writing – Original Draft, Visualization;

**Jeffrey C. Glennon:** Conceptualization, Funding acquisition, Methodology, Writing – review & editing

**Baziel G.M. van Engelen:** Conceptualization, Funding acquisition, Methodology, Resources, Supervision, Writing – review & editing

**Peter A.C. ‘t Hoen:** Conceptualization, Funding acquisition, Methodology, Supervision, Writing – review & editing

