## Supplemental Material for "Clinical improvement of DM1 patients reflected by reversal of disease-induced gene expression in blood"

**Supplementary Table 1: PCR Duplicates**

| PCR Sample ID | Patient ID | Reads in | Reads out | % Duplicates |
|---|---|---|---|---|
| 3 | D001_V4 | 74255337 | 6717608 | 91 |
| 4 | B053_V2 | 34902678 | 17657186 | 49 |
| 5 | B001_V2 | 32086534 | 19022483 | 41 |
| 6 | D001_V2 | 37216982 | 22233948 | 40 |
| 7 | C048_V4 | 97744825 | 6093127 | 94 |
| 9 | C055_V4 | 26857293 | 4802757 | 82 |
| 10 | B009_V4 | 27127348 | 5821860 | 79 |
| 12 | A005_V2 | 52839750 | 27213328 | 48 |
| 13 | B052_V4 | 46241711 | 9651283 | 79 |
| 14 | B028_V4 | 35160942 | 21099665 | 40 |
| 15 | B001_V4 | 55901222 | 25549654 | 54 |
| 16 | B042_V2 | 30533502 | 16419733 | 46 |
| 17 | B032_V2 | 38685827 | 12594622 | 67 |
| 18 | A017_V2 | 31667720 | 22563298 | 29 |
| 19 | B053_V4 | 34669594 | 22737682 | 34 |
| 20 | B011_V4 | 43683175 | 28303487 | 35 |
| 21 | D003_V4 | 29746212 | 9117004 | 69 |
| 23 | C048_V2 | 40956378 | 14674589 | 64 |
| 24 | C036_V2 | 31094075 | 18619186 | 40 |
| 25 | C039_V2 | 33732128 | 19923385 | 41 |
| 26 | C023_V4 | 87472685 | 8368215 | 90 |
| 27 | C039_V4 | 32324685 | 15233522 | 53 |
| 28 | C036_V4 | 29116832 | 6358863 | 78 |
| 29 | D037_V2 | 50853995 | 9424147 | 81 |
| 30 | D043_V2 | 38565222 | 9848583 | 74 |
| 31 | A023_V2 | 43568775 | 9619931 | 78 |
| 32 | A064_V2 | 52385485 | 9710934 | 81 |
| 33 | B032_V4 | 33748110 | 14557887 | 57 |
| 34 | C014_V4 | 64527362 | 8321368 | 87 |
| 35 | C004_V4 | 45992074 | 21544600 | 53 |
| 36 | C034_V4 | 33772304 | 20752983 | 39 |
| 37 | C014_V2 | 91287051 | 25791002 | 72 |
| 38 | C023_V2 | 32097203 | 9338738 | 71 |
| 39 | C019_V2 | 68218732 | 11297909 | 83 |
| 40 | A005_V4 | 37127933 | 16984404 | 54 |
| 41 | A038_V4 | 32282945 | 16454541 | 49 |
| 42 | A038_V2 | 73595623 | 8512630 | 88 |
| 43 | C004_V2 | 47664760 | 9628878 | 80 |
| 44 | C046_V2 | 32477430 | 16013060 | 51 |
| 45 | C034_V2 | 73271016 | 15781730 | 78 |
| 46 | C055_V2 | 39645604 | 10658203 | 73 |
| 47 | C046_V4 | 70648537 | 7596368 | 89 |
| 48 | D043_V4 | 85026900 | 6356019 | 93 |
| 49 | A064_V4 | 56851191 | 20306373 | 64 |

| | | | | |
|---|---|---|---|---|
| 50 | D037_V4 | 33753201 | 23851356 | 29 |
| 51 | A017_V4 | 31416730 | 17668191 | 44 |
| 53 | B052_V2 | 54599613 | 12969826 | 76 |
| 54 | B028_V2 | 75294410 | 7142975 | 91 |
| 55 | A023_V4 | 61933892 | 8854582 | 86 |
| 56 | B042_V4 | 44165589 | 13127682 | 70 |
| 57 | C019_V4 | 42917480 | 9427079 | 78 |
| 58 | B011_V2 | 30187435 | 12024301 | 60 |
| 59 | B009_V2 | 78351372 | 7813368 | 90 |
| 60 | D003_V2 | 42353399 | 18407056 | 57 |

For all the samples included in our analyses, table S1 illustrates the raw PCR counts as well as the percentage of PCR duplicates. Sampling timepoints during the trial can be derived from the Patient ID’s, with respectively V2 and V4 referring to before and after the intervention.

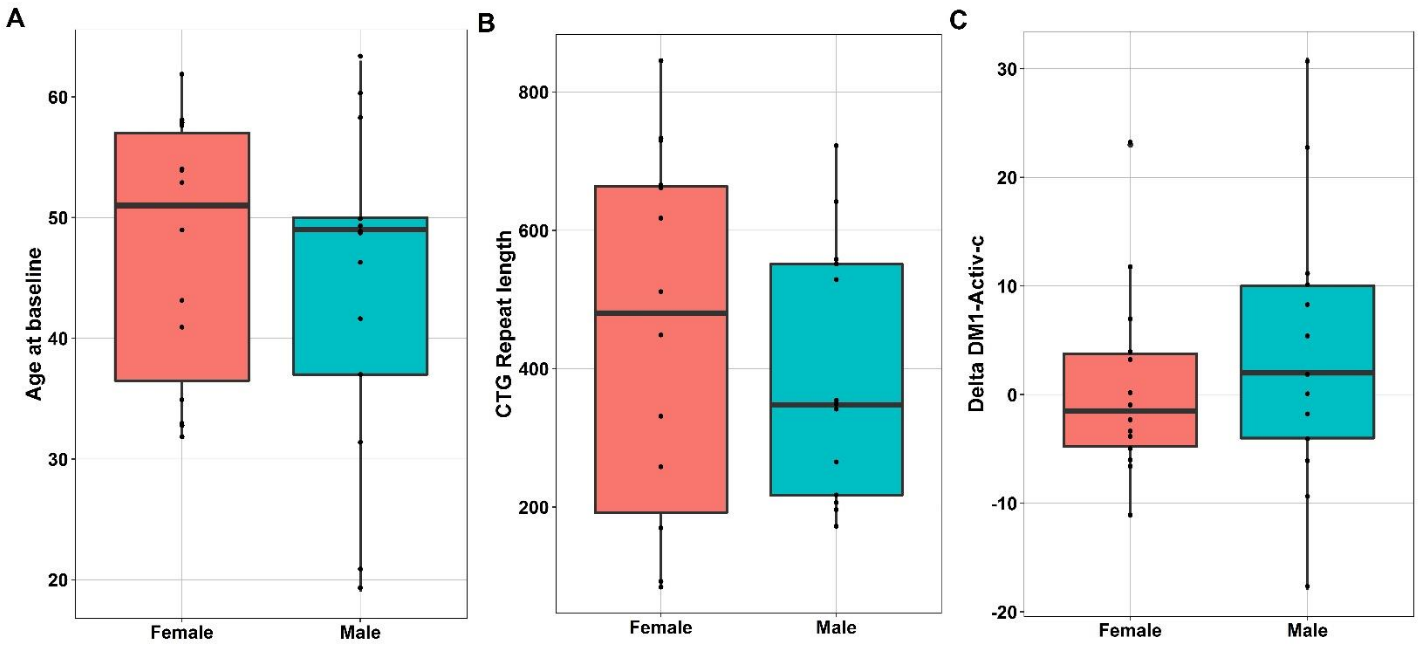


**Supplemental Figure S1: Patient characteristics**

Box-plots illustrating the distribution of age (A) and CTG-repeat length (B) of the 27 patients at baseline, separated by sex. C) Box-plots illustrating the change in DM1-Activ-c score after the intervention versus before, as expressed in Delta DM1-Activ-c scores, separated by sex.

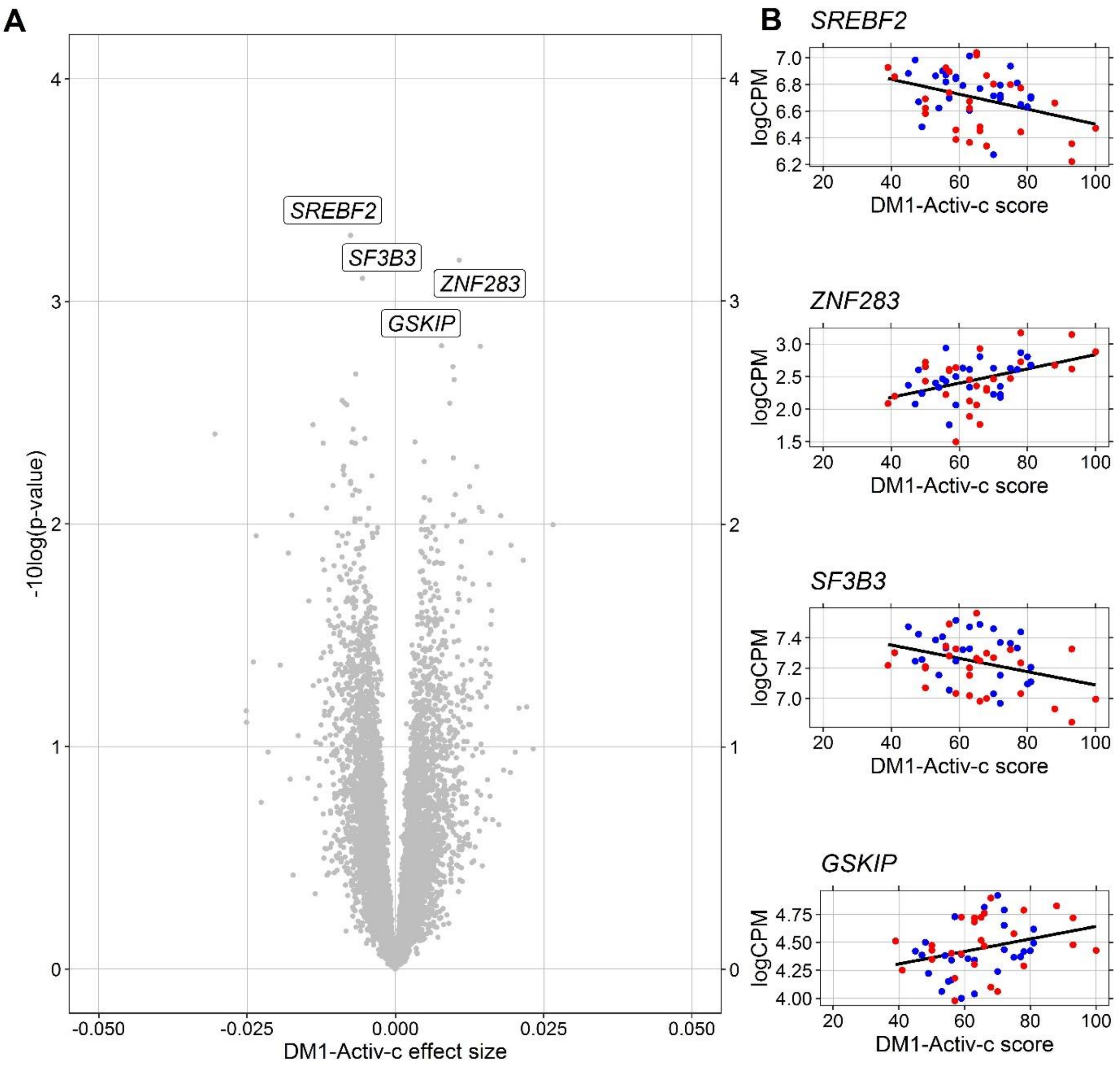


**Supplemental Figure S2: Gene expression association with DM1-Activ-c**

For each gene a mixed effect model was fitted with before/after CBT and DM1-Activ-c scores as fixed effects, while accounting for (random) effects of the individual. The p-values for the fixed effects were estimated via Satterthwaite's freedom method and FDR corrected. A) Volcano plot of the significance (-10log of the nominal p-value) and effect size of the DM1-Activ-c scores on gene expression. B) For the four genes with the lowest nominal p-values from panel A, the DM1-Activ-c scores are plotted against the gene expression values (logCPM). Blue dots represent baseline values, red dots values after CBT. The regression line indicates the linear association independent of the timepoints.

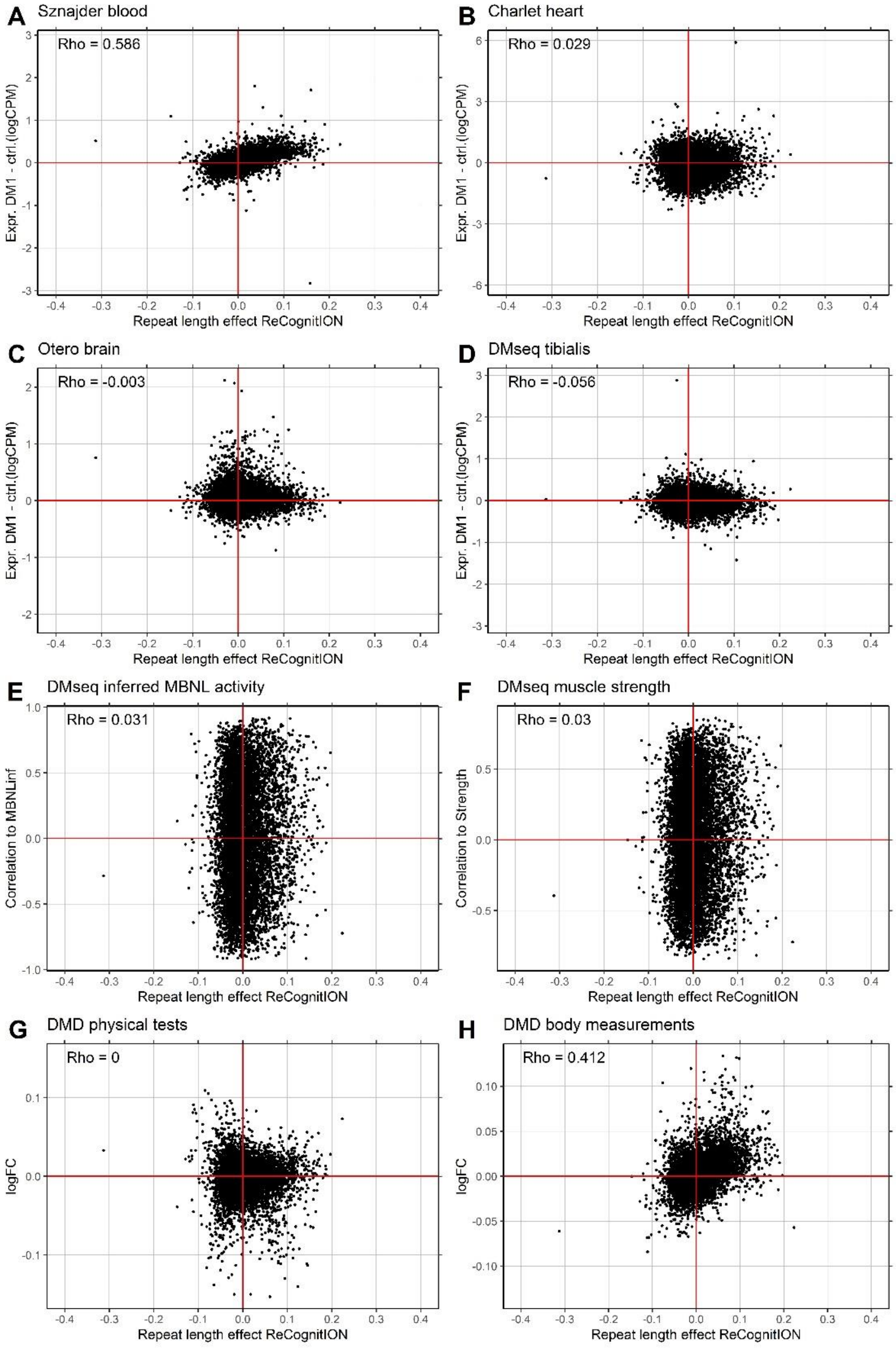


**Supplemental Figure S3. Comparison of gene expression associated to DM1 in other studies with that to the CTG repeat in this study**

The mean differences in normalized gene counts were calculated between DM1 and control samples for four studies comparing blood (A, [25]), heart (B, [15]), brain (C, [12]) and tibialis muscle (D, [11]) and plotted against the effect sizes for the CTG-repeat in this study for genes that were measured in both studies. In E and F, the correlation of expression to the inferred MBNL activity and muscle strength[11], was compared to our effects for the CTG-repeat. In G and H, gene expression associations in blood of the results of physical tests and several body measurements for Duchenne muscular dystrophy (DMD) patients [22] are compared to CTG-repeat associations from our study. In [22], associations with physical tests and body measurements were based on the first principal component over associated measures, each reflecting respectively 78% and 70% of the associated measurements variances. Depicted on the top left in each graph is the Pearson correlation coefficient for the plotted values.

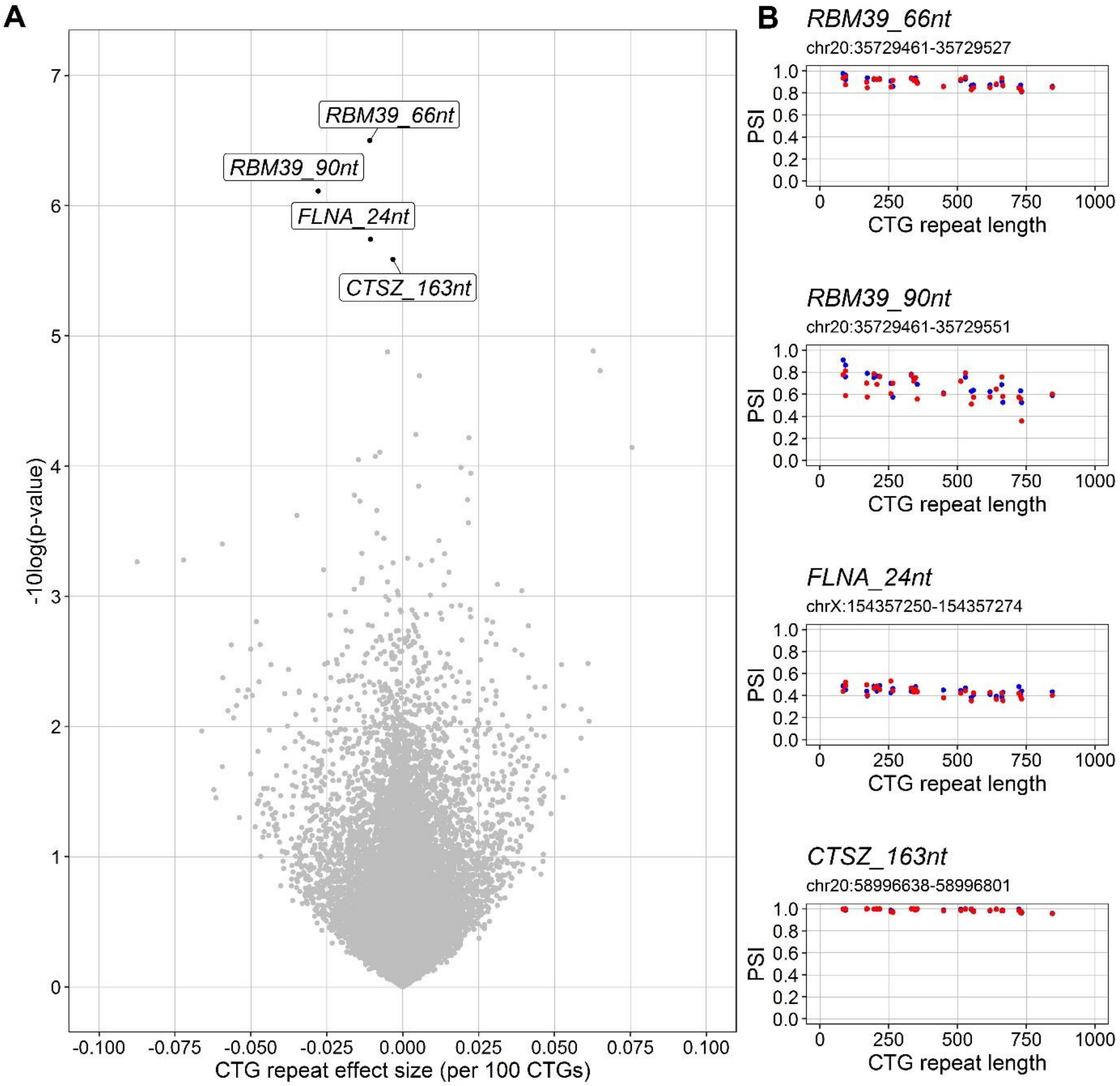


**Supplemental Figure S4. Splice exclusion and CTG-repeat length**

PSI values for splice exclusion events were determined using rMATS [. For each of the PSI values a linear mixed effect model was fitted with the modal CTG repeat length as covariate and patient as random effect. A) Volcano plot of significance (-log 10 of the nominal p-values of the modal CTG effect size) and the CTG effect size for the PSI values. Significant results after FDR correction ($p < 0.05$) are marked in black. B) For the four PSI values with the lowest nominal p-values from A, the PSI values are plotted against the modal CTG repeat lengths before (blue) and after the CBT intervention (red).

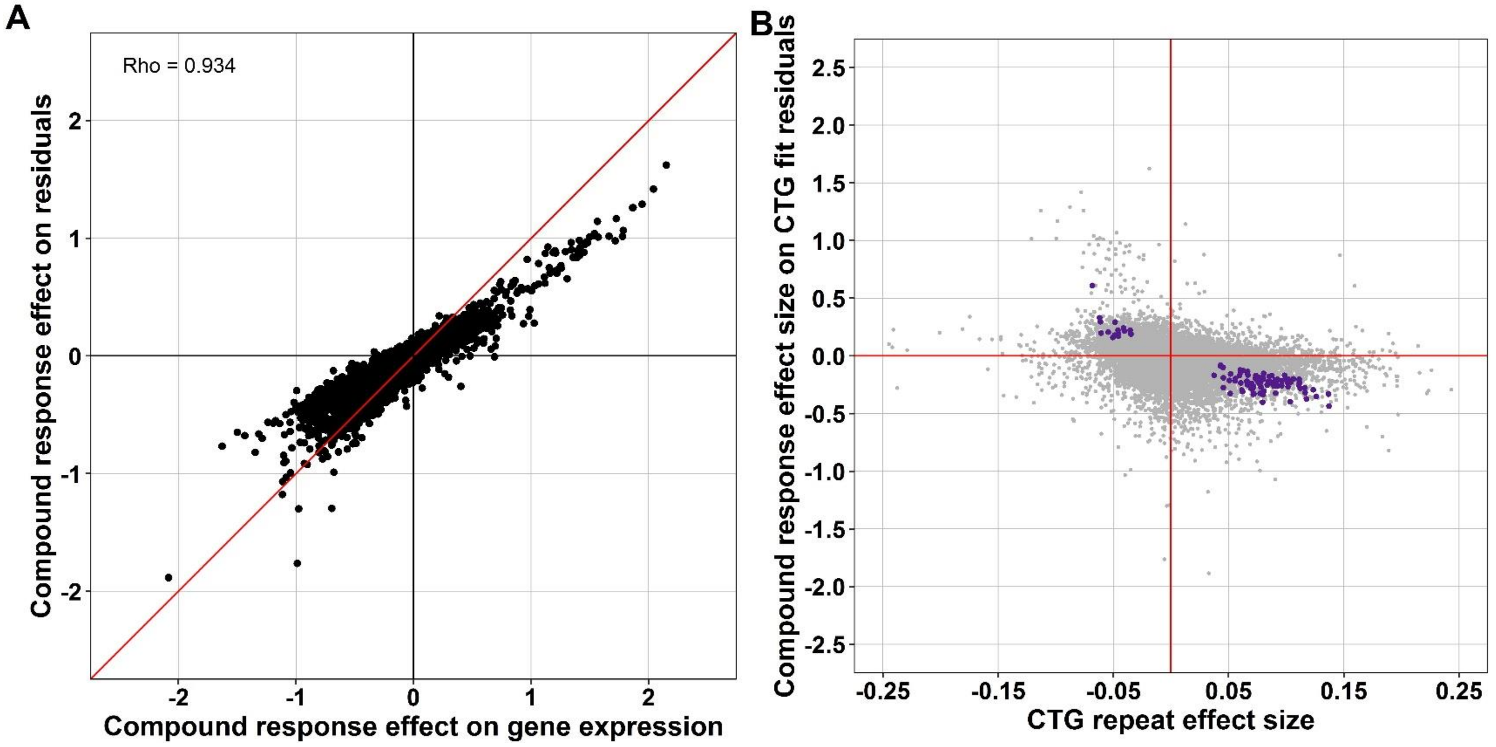


**Supplemental Figure S5. Shared explained variance among CTG-repeat and Compound Response predictors**

To assess the overlap in gene expression level variance explained by the CTG-repeat length and the Compound Response score, the Compound Response score was fitted on the residuals of the CTG-repeat length as fixed effect, while accounting for random effects of the patient. A) The effect sizes of the Compound Response score as estimated on the CTG-repeat model residuals are plotted against the effect sizes Compound Response scores as presented in this study. The Rho score reflects the Pearson correlation coefficient. B) Analogous to figure 5B, the Compound Response score effect size as estimated on the CTG-repeat model residuals are plotted against the CTG repeat effect size (per 100 CTG's). For illustrative purposes the x-axis has been scaled between -0.25 and 0.25, resulting in the removal of 12 outliers.
